# Comparator supplement patterns qualify the clinical interpretation of an EHR-derived glucosamine signal in Alzheimer’s disease

**DOI:** 10.64898/2026.07.23.26358748

**Authors:** Saki Nakashima, Kenichiro Sato, Yoshiki Niimi, Wataru Satake, Takeshi Iwatsubo

## Abstract

Electronic health records linked documented glucosamine use to faster progression from mild cognitive impairment (MCI) to Alzheimer’s disease (AD) and poorer survival after dementia. We examined these associations in the National Alzheimer’s Coordinating Center cohort and compared glucosamine with other supplement records captured in the same medication fields. The estimate for progression to primary AD dementia was in the same risk direction as the original finding, but similar estimates were observed for multivitamin/broad vitamin and calcium/vitamin D records. Among participants with dementia, glucosamine records were not associated with higher adjusted mortality, although death-data limitations reduce certainty. These findings do not address the experimental mechanism, but show that a glucosamine record alone does not establish a glucosamine-specific clinical effect.

## Introduction

Hawkinson et al. reported experimental evidence that hyperglycosylation may drive Alzheimer’s disease (AD) pathology and that oral glucosamine worsened social memory in 5xFAD mice.^1^ Their electronic health record (EHR) analysis also associated documented glucosamine use with progression from mild cognitive impairment (MCI) to AD and poorer survival in AD-related dementia.^1^ Previous population-based studies in cognitively unselected or dementia-free cohorts reported null or inverse associations with incident dementia but did not address the same clinical stages or outcomes.^2^–□ These experimental and observational findings raise different inferential questions. Stage specificity does not establish exposure specificity: the unresolved question is whether documented glucosamine use identifies a glucosamine-specific clinical signal in observational data. Because glucosamine is used mainly for osteoarthritis and joint symptoms, its documentation may also reflect indication, co-supplement use, healthcare contact, caregiver reporting and medication reconciliation. □–^11^

## Methods

We used the National Alzheimer’s Coordinating Center Uniform Data Set (NACC UDS)^12^–^1^□ as an external specificity assessment rather than a direct replication of the EHR design. NACC differs in recruitment, medication and outcome ascertainment, and follow-up structure. Glucosamine and comparator records were identified from the same medication-name fields. We prioritized

MCI-to-primary AD dementia and all-cause mortality among participants with dementia as the closest available counterparts to the original analyses. MCI-to-all-cause dementia, AD-associated dementia and normal cognition-to-MCI were retained as contextual or sensitivity endpoints (Supplementary Fig. 1 and Supplementary Tables 1 and 2). Comparators were used to assess whether the glucosamine pattern was distinctive within the same ascertainment system, not as strict negative controls.

## Results

Fully adjusted HRs for a baseline glucosamine record were 1.066 (95% CI, 0.917–1.239) for all-cause dementia, 1.109 (0.940–1.308) for AD-associated dementia and 1.155 (0.977–1.366) for primary AD dementia (Fig. 1a,b). The primary AD estimate was similar after ADL/IADL adjustment (HR, 1.177; 95% CI, 0.995–1.393; Supplementary Table 2). Although directionally compatible with the original EHR finding, it was not distinctive: multivitamin/broad vitamin and calcium/vitamin D records showed similar estimates (Fig. 1a and Supplementary Table 3). These exploratory comparisons do not establish effects of the comparator supplements, but they did not isolate a glucosamine-specific pattern.

**Figure 1.**
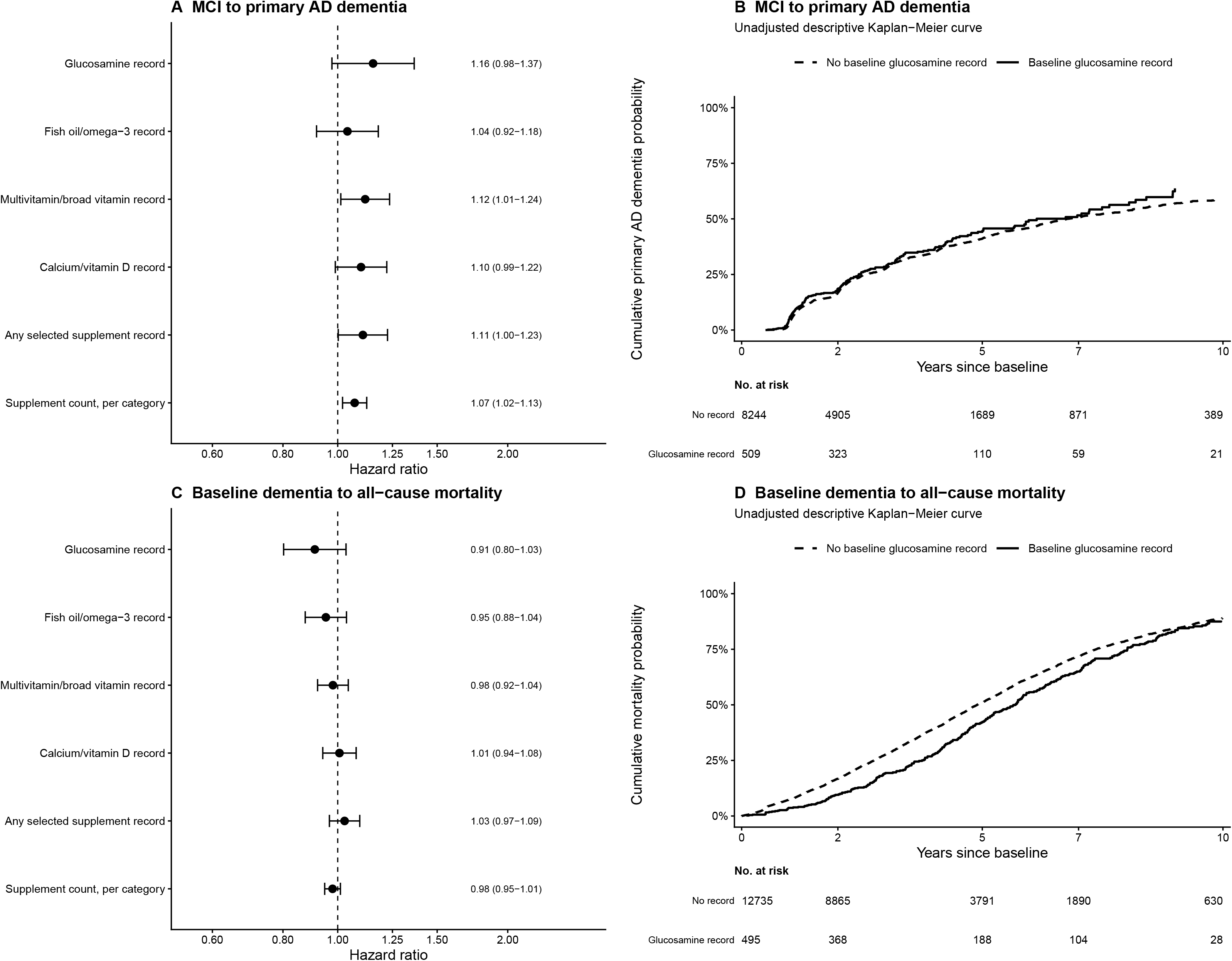
Recorded glucosamine and comparator supplement associations for the NACC outcomes most closely aligned with the original EHR analyses. **a**, Associations of baseline glucosamine and comparator supplement records with transition from mild cognitive impairment (MCI) to primary Alzheimer’s disease (AD) dementia. Points and horizontal lines show hazard ratios and 95% confidence intervals from separate exposure-specific, fully adjusted Cox proportional hazards models. Primary AD dementia was defined as incident dementia with AD recorded as the primary etiologic diagnosis; incident dementia not meeting this definition was treated as a censoring event. **b**, Unadjusted descriptive 1 − Kaplan–Meier curves for transition from MCI to primary AD dementia according to the presence or absence of a baseline glucosamine record. Numbers at risk are shown at 0, 2, 5, 7 and 10 years. **c**, Associations of baseline glucosamine and comparator supplement records with all-cause mortality among participants with dementia at baseline. Points and horizontal lines show hazard ratios and 95% confidence intervals from separate exposure-specific, fully adjusted clinical Cox proportional hazards models. **d**, Unadjusted descriptive Kaplan–Meier cumulative mortality curves according to the presence or absence of a baseline glucosamine record. Numbers at risk are shown at 0, 2, 5, 7 and 10 years. Fully adjusted models included age, sex, education, total medication count, baseline Clinical Dementia Rating–Sum of Boxes, stratification by Alzheimer’s Disease Research Center site, and available APOE ε4, vascular-history and medication-class covariates with less than 30% missingness. “Any selected supplement record” indicates at least one baseline record for glucosamine, chondroitin only, fish oil/omega-3, multivitamin/broad vitamin, or calcium/vitamin D. The hazard ratio for supplement count represents the association per additional recorded supplement category. Comparator analyses were exploratory and were not adjusted for multiple comparisons. Kaplan–Meier curves are unadjusted and are presented descriptively. For mortality, exact death dates were unavailable; deaths were assigned to 1 July of the reported year, and deaths occurring in the baseline calendar year were assigned 0.5 years of follow-up. The proportional-hazards assumption was not satisfied for the glucosamine term in the fully adjusted mortality model; mortality hazard ratios should therefore be interpreted as average summary estimates over follow-up and not as constant effects. The mortality estimates and unadjusted curves should not be interpreted as evidence of a survival benefit.

In the contextual normal cognition-to-MCI analysis, glucosamine and most comparator measures showed inverse estimates (Supplementary Fig. 1 and Supplementary Tables 1 and 2). We do not interpret these as evidence of benefit; the shared pattern is more compatible with supplement-user or ascertainment effects than compound-specific protection.

Among 13,230 participants with dementia at baseline, 8,414 died; 495 had glucosamine records, of whom 314 died (Fig. 1c,d). Glucosamine records did not support a higher mortality hazard after adjustment for age, sex, education, medication count, ADRC site and baseline CDR-SB (HR, 0.930; 95% CI, 0.826–1.046), or in the complete-case fully adjusted model (N = 10,462; 6,658 deaths; HR, 0.911; 95% CI, 0.802–1.034; Supplementary Table 4). We do not interpret these estimates as evidence of survival benefit. Death dates were year-based approximations, and proportional-hazards diagnostics indicated non-proportionality. Given these limitations, the result should be interpreted as discordant with, rather than as a direct replication test of, the original mortality association.

## Discussion

Comparator records provided a within-ascertainment check on specificity, but were not strict negative controls because their cognitive or survival effects cannot be assumed to be null.^1^□ The relevant question was whether glucosamine appeared distinctive from records captured through the same process. It did not for MCI-to-primary AD dementia, and several supplement categories also showed similar descriptive mortality patterns (Fig. 1 and Supplementary Tables 3 and 4). These exploratory analyses were not adjusted for multiple comparisons and do not establish effects of individual supplements.

We interpret NACC as a specificity assessment of the observational exposure record, not as evidence that glucosamine is harmless or as a test of the experimental mechanism. A stage-dependent biological effect remains possible, but shared patterns across supplement categories are compatible with residual confounding or ascertainment processes, including indication, healthcare contact, caregiver reporting and medication reconciliation. These factors were not measured directly. The narrower observation is that a glucosamine record did not behave as a uniquely harmful signal within the same ascertainment system.

NACC medication records do not capture over-the-counter dose, duration, adherence or formulation changes, and baseline records may be less precise for sustained exposure than EHR documentation. Our inference relies on within-system comparison, not on NACC ascertainment being superior. Participants are not population-representative, follow-up depends on continued participation and centre practices, and musculoskeletal indications were incompletely captured. The mortality analysis used all-cause dementia and death, year-based death dates and a model with non-proportional hazards. All estimates remain vulnerable to residual confounding, selection and exposure misclassification.

These findings do not challenge the experimental evidence that glucosamine can affect AD-relevant glycan biology. In NACC, the MCI-to-primary AD dementia estimate was in the same direction as the original EHR finding but was not distinctive from other supplement records, while the mortality analysis did not support a higher hazard. Documented glucosamine use should therefore not be assumed to identify a glucosamine-specific causal effect. A clinical trial should verify formulation, dose, duration, adherence, cognitive stage, indication and concomitant supplement use.

## Acknowledgements

We thank the participants, study partners, investigators and staff of the National Alzheimer’s Coordinating Center and the Alzheimer’s Disease Research Centers for making these data available. The NACC database is funded by NIA/NIH Grant U24 AG072122. NACC data are contributed by the NIA-funded ADRCs: P30 AG062429 (PI James Brewer, MD, PhD), P30 AG066468 (PI Oscar Lopez, MD), P30 AG062421 (PI Teresa Gomez-Isla, MD), P30 AG066509 (PI Thomas Grabowski, MD), P30 AG066514 (PI Mary Sano, PhD), P30 AG066530 (PI Helena Chui, MD, Arthur Toga, PhD), P30 AG066507 (PI Marilyn Albert, PhD), P30 AG066444 (PI David Holtzman, MD), P30 AG066518 (PIs Lisa Silbert, MD, Kevin Duff, PhD), P30 AG066512 (PI Thomas Wisniewski, MD), P30 AG066462 (PI Scott Small, MD), P30 AG072979 (PI David Wolk, MD), P30 AG072972 (Pis Charles DeCarli, MD, Rachel Whitmer, PhD), P30 AG072976 (PI Andrew Saykin, PsyD), P30 AG072975 (PI Julie Schneider, MD, MS), P30 AG072978 (PI Ann McKee, MD), P30 AG072977 (PI Robert Vassar, PhD), P30 AG066519 (PI Joshua Grill, PhD), P30 AG062677 (PIs Brad Boeve, MD, Ronald Petersen, MD, PhD), P30 AG079280 (PI Jessica Langbaum, PhD), P30 AG062422 (PI Gil Rabinovici, MD), P30 AG066511 (PI Allan Levey, MD, PhD), P30 AG072946 (PI Linda Van Eldik, PhD), P30 AG062715 (PI Sanjay Asthana, MD, FRCP), P30 AG072973 (PI Russell Swerdlow, MD), P30 AG066506 (PIs Glenn Smith, PhD, ABPP, David Lowenstein, PhD, Ranjan Duara, MD), P30 AG066508 (PIs Stephen Strittmatter, MD, PhD, Christopher Van Dyck, MD), P30 AG066515 (PI Victor Henderson, MD, MS), P30 AG072947 (PI Suzanne Craft, PhD), P30 AG072931 (PI Henry Paulson, MD, PhD), P30 AG066546 (PIs Sudha Seshadri, MD, Gladys Maestre, MD, PhD), P30 AG086401 (PI Erik Roberson, MD, PhD), P30 AG086404 (PI Gary Rosenberg, MD), P30 AG086403 (PI Angela Jefferson, PhD), P30 AG072958 (PIs Heather Whitson, MD, Gwenn Garden, MD, PhD), P30 AG072959 (PI Jagan Pillai, MD, PhD), P30 AG092752 (Ihab Hajjar, MD, MS).

## Author contributions

S.N. and K.S. conceived the comment, performed the analysis, and drafted the manuscript. Y.N., W.S. and T.I. provided clinical and methodological input. All authors reviewed and approved the manuscript.

## Competing interests

SN has no competing interests to disclose.

KS has no competing interests related to the content of the manuscript, is involved in a joint research project with the MetLife Foundation, and had received a research grant from Eli Lilly for collaborative research unrelated to the current manuscript.

YN has no conflicts of interest related to the content of the manuscript, is involved in collaborative researches with NIPRO Corporation, CANON Medical Systems Corporation, and Eli Lilly & Company, and had received consultancy/speaker fees from Eisai, and Eli Lilly.

TI has no conflicts of interest related to the content of the manuscript, had received consultancy/speaker fee from Biogen, Eisai, Eli-Lilly, and Roche/Chugai.

This manuscript has been prepared in a neutral and objective manner, and all disclosed financial relationships are not relevant to the content of this work.

## Ethical considerations

This secondary analysis used de-identified, controlled-access NACC UDS data obtained under the NACC Data Use Agreement. The study was approved by the University of Tokyo Graduate School of Medicine institutional ethics committee (ID: 2025264NI). The committee determined that additional informed consent was not required for this secondary analysis.

## Data availability

The data used in this study are available from the National Alzheimer’s Coordinating Center subject to data-use approval and the applicable NACC data-use agreement. The analytic datasets generated for this study are not publicly redistributed because they contain NACC-derived participant-level data.

## Code availability

Analysis code used to generate the reported estimates and figures will be made available upon reasonable request, subject to the terms of the NACC data-use agreement.

## Abbreviations

AD: Alzheimer’s disease
ADRC: Alzheimer’s Disease Research Center
APOE: apolipoprotein E
CDR-SB: Clinical Dementia Rating–Sum of Boxes
CI: confidence interval
EHR: electronic health record
HR: hazard ratio
MCI: mild cognitive impairment
NACC: National Alzheimer’s Coordinating Center.8

