## Supplementary Materials for "Comparator supplement patterns qualify the clinical interpretation of an EHR-derived glucosamine signal in Alzheimer’s disease"

**Supplementary Methods**

*Study population*

This analysis used the National Alzheimer’s Coordinating Center Uniform Data Set (NACC UDS), a multicentre observational research cohort of participants evaluated at Alzheimer’s Disease Research Centers.¹–³ Participants were included if they had baseline medication information and at least one follow-up visit. Separate baseline cohorts were constructed according to baseline cognitive diagnosis. The cognitively normal cohort was followed for incident MCI, the MCI cohort was followed for incident dementia outcomes, and the baseline dementia cohort was followed for all-cause mortality. The analysis was designed as an external specificity assessment of recorded glucosamine use rather than a direct replication of the original EHR design. The two outcomes closest to the original human analyses—MCI-to-primary AD dementia and baseline dementia-to-all-cause mortality—were emphasised in the main text; normal cognition-to-MCI and MCI-to-all-cause dementia were analysed as contextual endpoints.

*Exposure definitions*

Baseline glucosamine exposure was operationalized from medication-name text fields at the baseline visit and is referred to throughout as a baseline glucosamine record. The primary definition included glucosamine, glucosamine/chondroitin combinations, glucosamine with methylsulfonylmethane (MSM), and prespecified glucosamine-containing brand-like terms. Chondroitin-only records were excluded from the primary definition and analysed separately. Comparator supplement records—chondroitin only, fish oil/omega-3, multivitamin/broad vitamin, and calcium/vitamin D—were derived from the same medication-name fields. Additional measures captured any selected supplement record and the number of selected supplement categories. These comparators were used to assess whether the glucosamine estimate was distinctive within the same ascertainment system. They were not treated as strict negative-control exposures because their cognitive or survival effects cannot be assumed to be null.⁴

*Outcome definitions*

The AD-focused progression endpoint emphasised in the main text was first observed transition from baseline MCI to primary AD dementia, defined as incident dementia with AD recorded as the primary etiologic diagnosis. MCI-to-AD-associated dementia, defined as incident dementia with AD recorded as an etiologic or contributing diagnosis, was retained as a sensitivity endpoint.

The mortality endpoint was defined as time from the baseline dementia visit to all-cause death or censoring at the last available follow-up information. Because exact death dates were not available in the analytic extract, the primary analysis used July 1 of the reported year of death. Deaths occurring in the same calendar year as the baseline visit were assigned 0.5 years of follow-up. Sensitivity analyses used January 1 and December 31 of the reported year of death.

The contextual progression endpoints were first observed transition from baseline normal cognition to MCI and first observed transition from baseline MCI to any dementia diagnosis. Participants who developed dementia before an observed MCI diagnosis in the normal cognition cohort were censored at the dementia visit. Follow-up time was calculated from baseline to the first observed endpoint or to the last available follow-up visit for censored participants.

*Statistical analysis*

Cox proportional hazards models were used to estimate hazard ratios and 95% confidence intervals. All P values were two-sided. Each participant contributed once to each endpoint-specific baseline cohort, with repeated follow-up visits used to ascertain outcomes. Kaplan–Meier curves were used only for unadjusted descriptive visualization. Each supplement category was evaluated in a separate exposure model using the endpoint-specific covariates described below. These comparisons were descriptive and were not formal tests of equivalence or differences between supplement-specific coefficients. Supplement categories were not mutually exclusive. Comparator analyses were exploratory, were not adjusted for multiple testing and were interpreted as pattern-based specificity assessments rather than as confirmatory tests of individual supplements.

For the normal cognition-to-MCI analysis, the primary adjusted model included age, sex, years of education, total baseline medication count and Alzheimer’s Disease Research Center (ADRC) site stratification. We avoided using additional clinical covariates that were not consistently applicable to cognitively normal participants. For the MCI-to-all-cause dementia and MCI-to-primary AD dementia analyses, the fully adjusted model included age, sex, years of education, total baseline medication count, baseline Clinical Dementia Rating (CDR) Sum of Boxes (CDR-SB), apolipoprotein E ε4 (*APOE-*ε4) carrier status, diabetes, hypertension, hypercholesterolemia, stroke history, nonsteroidal anti-inflammatory drug use, diabetes medication use, antihypertensive medication use, lipid-lowering medication use and ADRC site stratification.

Additional sensitivity models adjusted for available activities of daily living (ADL) and instrumental activities of daily living (IADL) function using the IADL function score in the MCI cohort. Arthritis-related complete-case models additionally adjusted for an arthritis indicator when available and were used as supportive sensitivity analyses to examine potential indication-related confounding.

For the baseline dementia-to-all-cause mortality analysis, the fully adjusted clinical model included age, sex, years of education, total baseline medication count, baseline CDR-SB, APOE4 carrier status, diabetes, hypertension, hypercholesterolemia, stroke history, nonsteroidal anti-inflammatory drug use, diabetes medication use, antihypertensive medication use, lipid-lowering medication use and ADRC site stratification. The proportional-hazards assumption was evaluated using Schoenfeld residual-based tests. Because proportional-hazards diagnostics suggested non-proportionality for the mortality endpoint, Cox estimates for mortality were interpreted cautiously. Kaplan-Meier-based unadjusted descriptive restricted mean survival time (RMST) differences at 3, 5 and 10 years were added as sensitivity analyses using bootstrap standard errors.

**Supplementary Results**

Unless otherwise specified, hazard ratios (HRs) were estimated using Cox proportional hazards models. Values in parentheses are 95% confidence intervals (CIs). Participants, n indicates the number of participants included in each model after application of eligibility criteria and complete-case covariate requirements. Events, n indicates the number of outcome events in the corresponding model. P values <0.001 are shown as <0.001. For mortality RMST analyses, estimates are Kaplan-Meier-based unadjusted descriptive differences; positive differences indicate longer restricted mean survival time among exposed participants.

**Supplementary Table 1 | Cohort sizes, events and follow-up according to baseline glucosamine record for each endpoint**

| **Endpoint** | **Baseline glucosamine record** | **Participants** | **Events** | **Person-years** | **Incidence per 100 person-years** | **Follow-up, median (IQR), years** | **Maximum follow-up, years** |
| --- | --- | --- | --- | --- | --- | --- | --- |
| **Normal cognition to MCI** | Absent | 16446 | 3422 | 89219.3 | 3.84 | 4.05 (2.01-7.70) | 20.21 |
|  | Present | 1252 | 241 | 7665.5 | 3.14 | 5.36 (2.36-8.73) | 18.90 |
| **MCI to all-cause dementia** | Absent | 8244 | 3437 | 27765.7 | 12.38 | 2.24 (1.23-4.24) | 19.30 |
|  | Present | 509 | 224 | 1745.6 | 12.83 | 2.41 (1.25-4.42) | 17.02 |
| **MCI to primary AD dementia** | Absent | 8244 | 2625 | 27765.7 | 9.45 | 2.24 (1.23-4.24) | 19.30 |
|  | Present | 509 | 183 | 1745.6 | 10.48 | 2.41 (1.25-4.42) | 17.02 |
| **Baseline dementia to all-cause mortality** | Absent | 12735 | 8100 | 50388.2 | 16.08 | 3.18 (1.64-5.51) | 19.66 |
|  | Present | 495 | 314 | 2228.1 | 14.09 | 3.98 (1.97-6.37) | 18.29 |

Separate endpoint-specific cohorts were constructed from participants in the National Alzheimer’s Coordinating Center Uniform Data Set who had baseline medication information and at least one follow-up visit. A baseline glucosamine record was defined as a glucosamine-containing entry in the baseline medication-name fields and does not establish formulation, dose, duration or adherence. Events represent the first observed transition to the specified outcome. For the normal cognition-to-MCI endpoint, participants who developed dementia before an observed MCI diagnosis were censored at the dementia visit. For AD-focused dementia endpoints, a dementia diagnosis that did not meet the specified AD attribution was treated as a censoring event. Incidence rates were calculated as events per 100 person-years. For mortality, death dates were approximated as 1 July of the reported year of death; deaths occurring in the baseline calendar year were assigned 0.5 years of follow-up.

**Abbreviations**: **AD**, Alzheimer’s disease; **IQR**, interquartile range; **MCI**, mild cognitive impairment; **NACC**, National Alzheimer’s Coordinating Center; **UDS**, Uniform Data Set.

**Supplementary Table 2 | Associations of baseline glucosamine records with cognitive progression endpoints in primary and sensitivity Cox models**

| **Panel** | **Endpoint** | **Model** | **HR (95% CI)** | **P value** | **N** | **Events** | **Participants with glucosamine record** | **Events among participants with glucosamine record** |
| --- | --- | --- | --- | --- | --- | --- | --- | --- |
| **A** | Normal cognition to MCI | Unadjusted | 0.821 (0.721-0.936) | 0.003 | 17698 | 3663 | 1252 | 241 |
|  |  | Primary adjusted | 0.762 (0.664-0.875) | <0.001 | 17616 | 3637 | 1250 | 241 |
| **B** | MCI to all-cause dementia | Unadjusted | 1.023 (0.894-1.171) | 0.743 | 8753 | 3661 | 509 | 224 |
|  |  | Adjusted for age, sex and education | 0.987 (0.862-1.130) | 0.847 | 8720 | 3645 | 507 | 224 |
|  |  | + medication count | 1.000 (0.869-1.150) | 0.997 | 8720 | 3645 | 507 | 224 |
|  |  | + ADRC-site stratification | 0.977 (0.848-1.126) | 0.748 | 8720 | 3645 | 507 | 224 |
|  |  | + baseline CDR-SB | 1.100 (0.954-1.268) | 0.189 | 8720 | 3645 | 507 | 224 |
|  |  | Fully adjusted | 1.066 (0.917-1.239) | 0.407 | 7388 | 3147 | 457 | 208 |
|  |  | Fully adjusted + ADL/IADL | 1.088 (0.935-1.266) | 0.277 | 7252 | 3097 | 452 | 206 |
|  |  | Fully adjusted + arthritis indicator | 0.990 (0.759-1.290) | 0.939 | 3113 | 1038 | 196 | 66 |
|  |  | Fully adjusted + ADL/IADL + arthritis indicator | 0.988 (0.757-1.289) | 0.929 | 3058 | 1024 | 195 | 66 |
|  |  | Medication-form version 2/3 sensitivity | 1.110 (0.956-1.288) | 0.171 | 6287 | 2488 | 482 | 209 |
|  |  | Medication-form version 2/3 + arthritis indicator | 0.960 (0.747-1.233) | 0.748 | 3671 | 1206 | 225 | 71 |
| **C** | MCI to AD-associated dementia | Fully adjusted | 1.109 (0.940-1.308) | 0.221 | 7388 | 2595 | 457 | 175 |
|  |  | Fully adjusted + ADL/IADL | 1.132 (0.959-1.336) | 0.143 | 7252 | 2552 | 452 | 173 |
| **D** | MCI to primary AD dementia | Fully adjusted | 1.155 (0.977-1.366) | 0.092 | 7388 | 2436 | 457 | 171 |
|  |  | Fully adjusted + ADL/IADL | 1.177 (0.995-1.393) | 0.058 | 7252 | 2399 | 452 | 169 |

Hazard ratios and 95% confidence intervals were estimated using Cox proportional hazards models. Panel A shows transition from normal cognition to MCI; Panel B, transition from MCI to all-cause dementia; Panel C, transition from MCI to AD-associated dementia; and Panel D, transition from MCI to primary AD dementia. The primary adjusted model for normal cognition-to-MCI included age, sex, education, total medication count and stratification by ADRC site. For MCI-to-all-cause dementia, sequential models added total medication count, ADRC-site stratification and baseline CDR-SB. Fully adjusted models included age, sex, education, total medication count, baseline CDR-SB, ADRC-site stratification, and available APOE ε4, vascular-history and medication-class covariates with less than 30% missingness. ADL/IADL sensitivity models additionally included the derived IADL function score. Arthritis-indicator models were complete-case sensitivity analyses that additionally adjusted for available arthritis history. Medication-form sensitivity analyses were restricted to UDS form versions 2 and 3. Sample sizes vary because models used complete cases for their respective covariates. All P values are two-sided Wald-test values. No adjustment was made for multiple comparisons.

**Abbreviations**: **AD**, Alzheimer’s disease; **ADL**, activities of daily living; **ADRC**, Alzheimer’s Disease Research Center; **APOE**, apolipoprotein E; **CDR-SB**, Clinical Dementia Rating–Sum of Boxes; **CI**, confidence interval; **HR**, hazard ratio; **IADL**, instrumental activities of daily living; **MCI**, mild cognitive impairment; **N**, number of participants; **UDS**, Uniform Data Set.

**Supplementary Table 3 | Associations of baseline glucosamine and comparator supplement records with cognitive progression endpoints**

| **Panel** | **Endpoint** | **Exposure** | **HR (95% CI)** | **P value** | **N** | **Events** | **Participants with exposure record** | **Events among participants with exposure record** | **Exposure-specific PH P value** |
| --- | --- | --- | --- | --- | --- | --- | --- | --- | --- |
| **A** | MCI to primary AD dementia | Glucosamine record | 1.155 (0.977-1.366) | 0.092 | 7388 | 2436 | 457 | 171 | 0.468 |
|  |  | Fish oil/omega-3 record | 1.040 (0.917-1.180) | 0.537 | 7388 | 2436 | 1093 | 366 | <0.001 |
|  |  | Multivitamin/broad vitamin record | 1.119 (1.013-1.235) | 0.027 | 7388 | 2436 | 2399 | 817 | 0.340 |
|  |  | Calcium/vitamin D record | 1.100 (0.990-1.222) | 0.076 | 7388 | 2436 | 2344 | 725 | 0.825 |
|  |  | Any selected supplement record | 1.108 (1.002-1.225) | 0.045 | 7388 | 2436 | 3645 | 1155 | 0.023 |
|  |  | Supplement count, per category | 1.071 (1.020-1.126) | 0.006 | 7388 | 2436 | - | - | 0.047 |
|  |  | Chondroitin-only record | 1.175 (0.777-1.778) | 0.445 | 7388 | 2436 | 53 | 25 | Not assessed |
| **B** | MCI to all-cause dementia | Glucosamine record | 1.066 (0.917-1.239) | 0.407 | 7388 | 3147 | 457 | 208 | 0.931 |
|  |  | Fish oil/omega-3 record | 1.009 (0.903-1.128) | 0.869 | 7388 | 3147 | 1093 | 458 | <0.001 |
|  |  | Multivitamin/broad vitamin record | 1.096 (1.005-1.195) | 0.039 | 7388 | 3147 | 2399 | 1061 | 0.286 |
|  |  | Calcium/vitamin D record | 1.068 (0.974-1.170) | 0.161 | 7388 | 3147 | 2344 | 938 | 0.703 |
|  |  | Any selected supplement record | 1.108 (1.015-1.209) | 0.021 | 7388 | 3147 | 3645 | 1518 | 0.037 |
|  |  | Supplement count, per category | 1.045 (1.000-1.091) | 0.048 | 7388 | 3147 | - | - | 0.053 |
|  |  | Chondroitin-only record | 0.976 (0.657-1.448) | 0.902 | 7388 | 3147 | 53 | 27 | Not assessed |
| **C** | Normal cognition to MCI | Glucosamine record | 0.762 (0.664-0.875) | <0.001 | 17616 | 3637 | 1250 | 241 | 0.255 |
|  |  | Fish oil/omega-3 record | 0.814 (0.733-0.903) | <0.001 | 17616 | 3637 | 2721 | 498 | 0.099 |
|  |  | Multivitamin/broad vitamin record | 0.857 (0.789-0.931) | <0.001 | 17616 | 3637 | 5766 | 1038 | <0.001 |
|  |  | Calcium/vitamin D record | 0.845 (0.775-0.921) | <0.001 | 17616 | 3637 | 5995 | 1038 | 0.005 |
|  |  | Any selected supplement record | 0.873 (0.804-0.948) | 0.001 | 17616 | 3637 | 8665 | 1539 | 0.002 |
|  |  | Supplement count, per category | 0.890 (0.857-0.925) | <0.001 | 17616 | 3637 | - | - | <0.001 |
|  |  | Chondroitin-only record | 0.954 (0.616-1.479) | 0.834 | 17616 | 3637 | 72 | 21 | Not assessed |

Each exposure was evaluated separately in an endpoint-specific adjusted Cox proportional hazards model. For Panels A and B, fully adjusted models included age, sex, education, total medication count, baseline CDR-SB, ADRC-site stratification, and available *APOE* ε4, vascular-history and medication-class covariates with less than 30% missingness. For Panel C, models included age, sex, education, total medication count and ADRC-site stratification.

“Any selected supplement record” indicates at least one baseline record for glucosamine, chondroitin only, fish oil/omega-3, multivitamin/broad vitamin, or calcium/vitamin D. The hazard ratio for supplement count represents the association per additional recorded supplement category; binary exposed-participant and exposed-event counts are therefore not applicable. Comparator supplements were exploratory within-ascertainment specificity measures and were not assumed to be causally null exposures. Similarity in the direction or magnitude of estimates does not establish equivalent effects of the individual supplements. Exposure-specific proportional-hazards P values were derived from Schoenfeld residual-based tests. These diagnostics were not assessed for the chondroitin-only progression models. All P values are two-sided. No adjustment was made for multiple comparisons.

**Abbreviations**: **AD**, Alzheimer’s disease; **ADRC**, Alzheimer’s Disease Research Center; **APOE**, apolipoprotein E; **CDR-SB**, Clinical Dementia Rating–Sum of Boxes; **CI**, confidence interval; **HR**, hazard ratio; **MCI**, mild cognitive impairment; **N**, number of participants; **PH**, proportional hazards.

**Supplementary Table 4 | Associations of baseline supplement records with all-cause mortality among participants with dementia at baseline**

(Panel A)

| **Exposure** | **Model** | **HR (95% CI)** | **P value** | **N** | **Deaths** | **Participants with exposure record** | **Deaths among participants with exposure record** |
| --- | --- | --- | --- | --- | --- | --- | --- |
| **Glucosamine record** | Unadjusted | 0.849 (0.757-0.952) | 0.005 | 13230 | 8414 | 495 | 314 |
|  | Adjusted for age, sex, education, medication count and ADRC site | 0.818 (0.727-0.919) | <0.001 | 13077 | 8300 | 488 | 310 |
|  | + baseline CDR-SB | 0.930 (0.826-1.046) | 0.226 | 13077 | 8300 | 488 | 310 |
|  | Fully adjusted clinical model | 0.911 (0.802-1.034) | 0.150 | 10462 | 6658 | 424 | 271 |
| **Fish oil/omega-3 record** | Fully adjusted clinical model | 0.953 (0.876-1.036) | 0.260 | 10462 | 6658 | 1265 | 739 |
| **Multivitamin/broad vitamin record** | Fully adjusted clinical model | 0.981 (0.921-1.044) | 0.541 | 10462 | 6658 | 3003 | 1857 |
| **Calcium/vitamin D record** | Fully adjusted clinical model | 1.007 (0.941-1.078) | 0.837 | 10462 | 6658 | 2682 | 1540 |
| **Any selected supplement record** | Fully adjusted clinical model | 1.028 (0.967-1.094) | 0.375 | 10462 | 6658 | 4477 | 2674 |
| **Supplement count, per category** | Fully adjusted clinical model | 0.979 (0.948-1.011) | 0.199 | 10462 | 6658 | - | - |
| **Chondroitin-only record** | Fully adjusted clinical model | 0.625 (0.431-0.907) | 0.013 | 10462 | 6658 | 52 | 30 |

(Panel B)

| **Assigned death date** | **HR (95% CI)** | **P value** | **N** | **Deaths** | **Exposure-specific PH P value** |
| --- | --- | --- | --- | --- | --- |
| **1 January** | 0.913 (0.804-1.036) | 0.159 | 10462 | 6658 | 0.030 |
| **1 July** | 0.911 (0.802-1.034) | 0.150 | 10462 | 6658 | 0.019 |
| **31 December** | 0.907 (0.798-1.030) | 0.132 | 10462 | 6658 | 0.016 |

(Panel C)

| **Exposure** | **Model** | **N** | **Deaths** | **Exposure-specific PH P value** | **Global PH P value** |
| --- | --- | --- | --- | --- | --- |
| **Glucosamine record** | Unadjusted | 13230 | 8414 | 0.001 | 0.001 |
|  | Adjusted for age, sex, education, medication count and ADRC site | 13077 | 8300 | <0.001 | <0.001 |
|  | + baseline CDR-SB | 13077 | 8300 | 0.001 | <0.001 |
|  | Fully adjusted clinical model | 10462 | 6658 | 0.019 | <0.001 |
| **Fish oil/omega-3 record** | Fully adjusted clinical model | 10462 | 6658 | 0.097 | <0.001 |
| **Multivitamin/broad vitamin record** | Fully adjusted clinical model | 10462 | 6658 | 0.081 | <0.001 |
| **Calcium/vitamin D record** | Fully adjusted clinical model | 10462 | 6658 | <0.001 | <0.001 |
| **Any selected supplement record** | Fully adjusted clinical model | 10462 | 6658 | <0.001 | <0.001 |
| **Supplement count, per category** | Fully adjusted clinical model | 10462 | 6658 | <0.001 | <0.001 |
| **Chondroitin-only record** | Fully adjusted clinical model | 10462 | 6658 | 0.468 | <0.001 |

(Panel D)

| **Exposure** | **Truncation time, years** | **Unexposed N** | **Exposed N** | **RMST, unexposed, years** | **RMST, exposed, years** | **Difference, exposed minus unexposed, years (95% CI)** | **P value** |
| --- | --- | --- | --- | --- | --- | --- | --- |
| **Glucosamine record** | 3 | 12735 | 495 | 2.622 | 2.792 | 0.170 (0.117-0.223) | <0.001 |
|  | 5 | 12735 | 495 | 3.835 | 4.211 | 0.376 (0.256-0.487) | <0.001 |
|  | 10 | 12735 | 495 | 5.152 | 5.766 | 0.614 (0.316-0.914) | <0.001 |
| **Fish oil/omega-3 record** | 3 | 11769 | 1461 | 2.612 | 2.763 | 0.151 (0.116-0.183) | <0.001 |
|  | 5 | 11769 | 1461 | 3.817 | 4.104 | 0.287 (0.216-0.369) | <0.001 |
|  | 10 | 11769 | 1461 | 5.122 | 5.616 | 0.494 (0.309-0.669) | <0.001 |
| **Multivitamin/broad vitamin record** | 3 | 9662 | 3568 | 2.599 | 2.707 | 0.107 (0.080-0.135) | <0.001 |
|  | 5 | 9662 | 3568 | 3.803 | 3.974 | 0.171 (0.109-0.230) | <0.001 |
|  | 10 | 9662 | 3568 | 5.122 | 5.321 | 0.199 (0.069-0.321) | 0.002 |
| **Calcium/vitamin D record** | 3 | 9979 | 3251 | 2.595 | 2.731 | 0.136 (0.107-0.162) | <0.001 |
|  | 5 | 9979 | 3251 | 3.794 | 4.019 | 0.225 (0.165-0.284) | <0.001 |
|  | 10 | 9979 | 3251 | 5.114 | 5.358 | 0.244 (0.123-0.377) | <0.001 |
| **Any selected supplement record** | 3 | 7855 | 5375 | 2.573 | 2.710 | 0.137 (0.114-0.164) | <0.001 |
|  | 5 | 7855 | 5375 | 3.758 | 3.983 | 0.225 (0.171-0.286) | <0.001 |
|  | 10 | 7855 | 5375 | 5.077 | 5.315 | 0.237 (0.117-0.362) | <0.001 |

All-cause mortality was evaluated among participants with dementia at baseline. A baseline supplement record denotes an entry in the baseline medication-name fields and does not establish formulation, dose, duration or adherence. Exact death dates were unavailable; the primary analysis assigned death to 1 July of the reported year, with deaths occurring in the baseline calendar year assigned 0.5 years of follow-up.

**Panel A** presents Cox proportional hazards estimates. For glucosamine, sequential models included an unadjusted model; adjustment for age, sex, education, total medication count and ADRC site; additional adjustment for baseline CDR-SB; and a fully adjusted clinical model. Fully adjusted clinical models included age, sex, education, total medication count, baseline CDR-SB, ADRC-site stratification, and available APOE ε4, vascular-history and medication-class covariates with less than 30% missingness. Comparator supplements were evaluated separately using the same fully adjusted clinical model. The hazard ratio for supplement count represents the association per additional recorded supplement category; binary exposed-participant and exposed-death counts are therefore not applicable.

**Panel B** presents sensitivity analyses assigning death to 1 January, 1 July or 31 December of the reported year.

**Panel C** presents exposure-specific and global Schoenfeld residual-based tests of the proportional-hazards assumption. Because proportional hazards were not satisfied for the glucosamine term in the fully adjusted mortality model, Cox hazard ratios should be interpreted as average summary estimates over follow-up rather than as constant effects.

**Panel D** presents unadjusted descriptive restricted mean survival time estimates truncated at 3, 5 and 10 years. Differences are calculated as the RMST among participants with an exposure record minus the RMST among those without that record. These unadjusted RMST estimates should not be interpreted as evidence of a survival benefit. Comparator analyses were exploratory, and no adjustment was made for multiple comparisons. All reported P values are two-sided.

**Abbreviations**: **ADRC**, Alzheimer’s Disease Research Center; **APOE**, apolipoprotein E; **CDR-SB**, Clinical Dementia Rating–Sum of Boxes; **CI**, confidence interval; **HR**, hazard ratio; **N**, number of participants; **PH**, proportional hazards; **RMST**, restricted mean survival time.

**Supplementary Figure 1 | Contextual analyses of recorded supplement associations with normal cognition-to-MCI and MCI-to-all-cause dementia transitions**

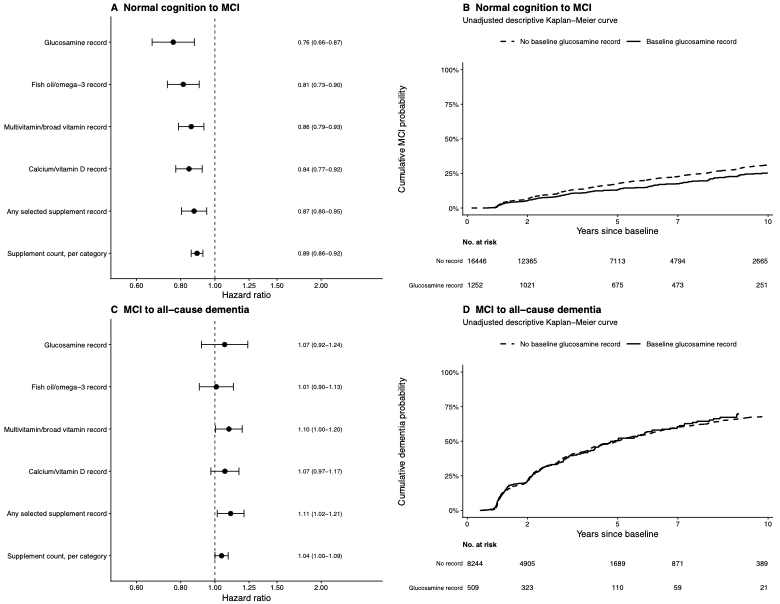

**a**, Associations of baseline glucosamine and comparator supplement records with transition from normal cognition to mild cognitive impairment (MCI). Points and horizontal lines show hazard ratios and 95% confidence intervals from separate exposure-specific adjusted Cox proportional hazards models. Participants who developed dementia before an observed MCI diagnosis were censored at the dementia visit.

**b**, Unadjusted descriptive Kaplan–Meier cumulative transition curves for normal cognition to MCI according to the presence or absence of a baseline glucosamine record. Numbers at risk are shown at 0, 2, 5, 7 and 10 years.

**c**, Associations of baseline glucosamine and comparator supplement records with transition from MCI to all-cause dementia. Points and horizontal lines show hazard ratios and 95% confidence intervals from separate exposure-specific, fully adjusted Cox proportional hazards models.

**d**, Unadjusted descriptive Kaplan–Meier cumulative transition curves for MCI to all-cause dementia according to the presence or absence of a baseline glucosamine record. Numbers at risk are shown at 0, 2, 5, 7 and 10 years.

The adjusted normal cognition-to-MCI models included age, sex, education, total medication count and stratification by Alzheimer’s Disease Research Center site. Fully adjusted MCI-to-all-cause dementia models included age, sex, education, total medication count, baseline Clinical Dementia Rating–Sum of Boxes, site stratification, and available APOE ε4, vascular-history and medication-class covariates with less than 30% missingness. “Any selected supplement record” indicates at least one baseline record for glucosamine, chondroitin only, fish oil/omega-3, multivitamin/broad vitamin, or calcium/vitamin D. The hazard ratio for supplement count represents the association per additional recorded supplement category.

Comparator analyses were exploratory and were not adjusted for multiple comparisons. Kaplan–Meier curves are unadjusted and are presented descriptively. These endpoints were analysed as contextual and bias-diagnostic analyses rather than as primary tests of the original AD-focused EHR findings. The lower normal cognition-to-MCI estimates should not be interpreted as evidence that glucosamine or the comparator supplements prevent cognitive decline.

**Abbreviations**: **AD**, Alzheimer’s disease; **ADRC**, Alzheimer’s Disease Research Center; **APOE**, apolipoprotein E; **CDR-SB**, Clinical Dementia Rating–Sum of Boxes; **CI**, confidence interval; **EHR**, electronic health record; **HR**, hazard ratio; **MCI**, mild cognitive impairment.
